# Quantitatively Evaluating the Cross-Sectoral and One Health Impact of Interventions: A Scoping Review and Application to Antibiotic Resistance

**DOI:** 10.1101/2020.01.30.20019703

**Authors:** Nichola R. Naylor, Jo Lines, Jeff Waage, Barbara Wieland, Gwenan M. Knight

## Abstract

Current published guidance on how to evaluate antibiotic resistance (ABR) from a One Health perspective has focussed on the evaluation of intervention design and of the intervention implementation process. For efficient resource allocation, it is also important to consider quantitative measures of intervention impact. In particular, there has been little discussion of how to practically evaluate ABR-related agri- and aquaculture interventions from a public health perspective. Lessons can be learned from other One Health and cross-sectoral intervention impact evaluations.

WebofScience, EconLit, PubMed and grey literature were searched for literature quantitatively evaluating interventions across humans, animals and/or the environment. The review included 90 studies: 73 individual evaluations (from 72 papers) and 18 reviews, all including some measure of human impact, but only 29 papers covered all three One Health perspectives (human, animal and environmental). To provide decision makers with expected outcome estimates that are related to their objective functions, evaluations should provide outcome estimates from different perspectives. These include individual, microeconomic and/or macroeconomic perspectives across the One Health system. Based on the methods found in this review, a multi-level compartmental modelling approach for ABR-related intervention evaluation is proposed. The outcomes of such models can then feed into multi-criteria-decision analyses that weigh respective impact estimates alongside other chosen outcome estimates (for example equity or uncertainty). It is key that future quantitative evaluation models of ABR-related interventions are shared (for example through open source code sharing websites) to avoid duplication of effort and to enable more comprehensive estimates of intervention impact to be modelled in the future.

## Introduction

Antibiotic resistance (ABR) may reduce our ability to prevent and treat bacterial infections in humans and in animals [1]. It has been described as a true One Health (OH) issue [2–4], in which OH can be defined as the description of and interactions between the individual levels of health, population levels of health and ecosystem levels of health (across humans, animals, plants and the wider environment) [5]. For the purposes of this paper, cross-sectoral relates to the interaction between two or more of these ecosystem factors (human, animal and ‘plants and environment’).

Policy options have been put forward to tackle further emergence and spread of ABR, through international policy reports and action plans [6, 7]. Taking antibiotic stewardship as an example, though there is some evidence of a positive effect of reduced food-producing animals’ antibiotic use on ABR outcomes in humans [8], there is a lack of evidence quantifying the wider socio-economic and OH impact of such interventions [9, 10]. Initiatives such as the Network for Evaluation of One Health (NEOH), alongside other available literature, offer frameworks for evaluation in OH topics, and introductions into methods of economic evaluations [11–13]. Such frameworks do mention the need for the quantitative evaluation of policy impact on outcomes. However, they do not propose a method to economically evaluate ABR from the perspectives of all stakeholders within the OH-system.

Quantitative (numerical) and qualitative (non-numerical) evaluations are needed to understand intervention implementation and impact. Given the complexity of integrating OH evaluations across decision makers representing different sectors, existing standard evaluation guidance or checklists do not offer practical discussions and applications for quantitative, impact evaluations across OH [14]. Such discussion on complex, OH intervention evaluation is needed to aid future evaluators in establishing their protocols and to indicate to policy/decision makers what information should be considered when deciding whether or not to update an intervention or fund an intervention evaluation.

It has previously been highlighted that ABR poses similar theoretical evaluation issues to climate change and zoonotic infection, due to their shared cross-sectoral and/or externality-inducing nature [15–17]. Impact evaluation literature estimating the effectiveness of interventions targeting zoonotic infections or climate change (cutting across the OH system) is available [15, 18]. Hence there is scope to learn from, and adapt, existing cross-sectoral evaluation approaches from these fields within the discipline of ABR-OH-economic evaluations. In order to do this, we first must understand what methods such evaluations have utilised, and discuss the appropriateness of these approaches within the context of ABR.

This review aimed to present a detailed discussion of impact evaluation perspectives, outcome measures and methods in relation to the quantitative evaluation of ABR-related interventions, utilising previous literature on cross-sectoral and OH interventions. In order to achieve this, the current paper aimed to (i) collate and describe previous methods used in the quantitative evaluation of interventions related to cross-sectoral and OH issues and (ii) utilise this information to construct a conceptual evaluative model for ABR-related interventions.

## Materials and Methods

### (i) Literature Review of One Health & Cross-Sectoral Intervention Evaluations

A rapid, scoping review method was utilised, enabling the targeting of specific areas of the literature that were hypothesised as important/resourceful by the authors. General OH evaluation literature was sought, but more specifically retrievals that were likely to provide a high inclusion rate (such as focusing on OH surveillance evaluation literature) and/or that were likely to be relevant to the case of ABR (such as “climate change” [17]) were wanted. The review was performed in WebofScience, EconLit, PubMed, Google databases [19–22], with 28^th^ March 2019 being the last search date (see Supplementary Material for search terms).

Titles were reviewed for appropriateness, followed by abstracts (if available), and subsequently full texts by the lead author. Where appropriate, the reference lists were also searched. A study had to include impact estimates across more than one sector (across human, animal and environment) within the intervention evaluation. If literature reviews were retrieved that offered results or conclusions that were directly relevant to this study they were included in the review results directly. Quantitative outcomes were defined as those with numerical valuations attached, and subsequently included. Only English language papers were included in this review, with no time period cut-off specified. For further description of inclusion/exclusion criteria applied see Supplementary Material Table 1.

**Table 1.** Scoping Review Indicator Definitions

| Indicator | Classification | Definition |
| --- | --- | --- |
| One Health Perspective | Human | Impact quantified on a person; including patients, farmers and individuals within the system under evaluation. |
|  | Animal | Impact quantified on animals, including livestock, fish, companion animals. |
|  | Environment | Impact quantified on the environment, including on temperature, water levels and on crops. |
| Evaluation Perspective | Individuals | Evaluating impact on mortality, morbidity, income at the individual level. |
|  | Microeconomic (Firm and Sector) | Evaluating impact within one specific sector; such as health care, environmental and agricultural sectors individually. It also included individual business impact, such as farm-level impact. |
|  | Macroeconomic (Multi-sector and Government) | Evaluating impact across multiple sectors within an economy or globally. |
| Methodology Perspective | Mathematical Simulation | Methods which take a hypothetical sample and model potential interactions and/or outcomes using mathematical formulae. This ranges from simple stepwise calculation methods (e.g. applying prevalence levels to a population of interest) to complex system dynamic models and general computable equilibrium models [24]. |
|  | Statistical Evaluation | Methods which take empirical data and apply statistical methods to estimate interactions and/or associated outcomes. This ranges from the calculation of descriptive statistics to complex survival analyses and regression analyses [24]. |
|  | Index/Rank Calculation | Methods which utilise a framework which compiles an index to measure the intervention. The two main indices included in this review are multi-criteria decision analysis (which creates weights and rankings across different decision criteria) and the One Health Index (which measures “One Healthness” [13]). |
|  | Other | If the study method did not fit into any of the above methodology perspective categories, then Other was used. |

Across included individual studies specific variables extracted were compared (such as temporal and geographical scope, a full list can be found in Supplementary Material Table 2) and suitable indicators created during data extraction. Indicators created for the purposes of this review were ‘One Health perspective’, ‘evaluation perspective’ and ‘methodology perspective’ (Table 1). Studies could cross multiple indicators (e.g. perform evaluations from both microeconomic and macroeconomic perspectives). A narrative synthesis of the included studies and their methods was performed. Given the expected heterogeneity in study design, and that no concluding statements were to be made about actual intervention effectiveness or efficiency, no formal quality assessment checklist was utilised.

**Table 2.** Potential Economic Agent Objectives and Constraints

| Agent | Potential Objectives and Constraints | Example(s) from the Literature |
| --- | --- | --- |
| Individuals | Individuals may seek to maximize individual expected utility over their lifetime (or over other pre-defined time horizons), based on savings, consumption of commodities and consumption of leisure, this includes the consumption of healthcare. This will be subject to budget constraints (a function of income). Broadening this, individuals could also seek to maximize individual capability [38]. | *Smith et al.* directly define a utility maximisation problem using equations involving utility values attributed to the consumption of commodities and leisure, solved using computable general equilibrium techniques [58]. |
| Firm | Firms may seek to maximise utility (a function of profits), based on the consumption of labour, capital and intermediate inputs, subject to the price of the inputs and output. However, risk or uncertainty may also factor into decision making processes of firms.<br>Time horizons, over which a firm's objective function is maximised, will depend on the nature of the production process and individual firms (for example this could be per quarter, financial year or crop cycle). | A review of farm-level evaluations found that half of the studies used profit maximization as the objective function, 29% included risk or stochasticity, whilst 18% of the studies used multi-criteria objective functions (including income, risk and labour factors) [41].<br>A study on water resource management adaptation policies discussed utility maximisation is often equated to profit maximisation for firms, however this study proposes using a multi-attribute utility maximization when using a firm perspective, with factors such as risk avoidance and management complexity included [59]. |
| Sector | These decision makers may be attempting to maximize the return on an investment within that sector. This could involve maximizing the expected health-related quality of life or monetary return of a given investment, or maximizing productivity (rate of such outputs for a given set of inputs), constrained by different financing issues depending on the sector and its context (e.g. public versus private). These factors will also impact desired time horizon for which the objective maximisation process is considered.<br>Increasing population capability could alternatively be the motivation, either through maximisation of total capability or through meeting a threshold level of capacity for as many people in society as possible [38]. | By using cost-utility analyses, such as estimating cost per disability-adjusted life year averted [60], studies assumed that payers want to maximise expected population utility, subject to budget constraints.<br>In cost-benefit analyses which apply a set monetary value to life/utility, there's an assumption that a positive return on investment is the goal of the intended [44]. |
| Government | A general objective function proposed for a nation's government is that of maximizing government utility from the consumption of commodities, capital and labour, constrained by tax revenues [58], and loans or other financing mechanisms. Government utility, however, could encompass wider social preferences in the form of being a function of equity, capability and risk/uncertainty.<br>Chosen time horizons and time preference assumptions may differ depending on policy perspectives and external forces (such as political election or budgetary cycles). | Previous reviews highlighted the importance of outcomes such as equity, capability, sustainability, uncertainty and animal welfare [39–42, 61]. A social cost-benefit analysis included various social discount rates alongside a survey on what the appropriate discount rate would be [62].<br>Economic agent objectives were incorporated into a climate change policy evaluation found in this review in the form of a 'multi-level model'. This allowed for an international level and lower levels to be linked and was based on a game of negotiations, using an underlying multi-attribute utility function [63]. One review found highlighted the lack of agricultural/food-security evaluations that included potentially important criteria such as economic development, environmental sustainability and peace [67]. |

### (ii) Evaluating Antibiotic-resistance Interventions

A hypothetical case study of conducting a quantitative evaluation for ABR interventions was chosen to build a conceptual evaluative model around. Antibiotic stewardship across agriculture and human healthcare systems was chosen, as this is highlighted as a key area for intervention by many of the AMR-policy documents and by nature is cross-sectoral [3, 23]. However, the key discussion points and recommendations are generalizable across different ABR-related interventions. A visual schematic of the conceptual evaluative model was drafted, with methodological recommendations narratively described. These include: perspectives (see Table 1), scope (temporal), model structure and basic model assumptions.

## Results

### (i) Literature Review of One Health & Cross-sectoral Intervention Evaluations

1479 unique retrievals followed by 170 full texts were screened from the formal searches, leading to 65 studies being included. Through reference lists and Google searches, an additional 25 papers were added. 72 studies (73 individual evaluations) underwent data extraction, whilst 18 reviews were included in the narrative synthesis. Individual study information can be found within Supplementary Material Tables 2 and 3. In terms of OH perspectives; all studies included a human impact (73 evaluations), with 84% including an environmental impact (61 evaluations) and 56% including an animal impact (41 evaluations). Only 29 (40%) of the 73 evaluations included all 3 perspectives.

#### Evaluation Perspective

In terms of evaluation perspective, 4 studies focused on individual perspectives, 42 had microeconomic perspectives and 51 included macroeconomic perspectives. The majority of studies only covered one of these evaluation perspectives (53%), with 40% having two evaluation perspectives (mainly covering microeconomic and macroeconomic) and 7% (5 studies) covering all three.

A number of the reviews offered a guide/framework to the approach of evaluation [13, 15, 25, 26], emphasising the importance of the following; (1) ensuring the definition of the research question is precise (including in terms of scope of interventions, comparators, risks assessed, system, stakeholders, scenarios and time), (2) defining inputs and outputs to the system (such as agents, actions and resources) and mapping out interactions between these, and (3) considering appropriate methods that capture the whole system that is impacted by changes through the intervention, and value outcomes accordingly.

The NEOH handbook provides an index to measure the “One Health-ness” of an intervention or issue (the OH index) [13], which had been applied in 8 individual evaluations included in this review [27–34]. The OH index evaluates the ‘degree and structural balance’ for OH-ness, including impact on OH thinking, planning and working [35]. Current applications of the OH index found are more process-focused (evaluating intervention design and implementation), rather than intervention impact focused.

Further literature found highlights that once stakeholders within a specified system have been identified, their decision making objectives and motives should be also be clearly outlined [36, 37]. One study stated that evaluations should be tailored to the decision making objectives of stakeholders, carefully linking monitoring and evaluation criteria to such objectives [36]. In Table 2, possible objectives of these different economic agents are presented, with relevant examples drawn from literature. The main economic theories used in the selection of potential objectives in Table 2 are grounded in utilitarianism (maximising overall ‘satisfaction’) as the main social justice perspective. However, the possibility to include egalitarian motives or Amartya Sen’s capability approaches was also noted [38]. Uncertainty was also highlighted in previous reviews as something that should be explicitly considered and quantified, as also valued by decision makers [36, 39–41].

#### Methodology Perspective

Reviews included in this narrative synthesis (summarised in Supplementary Material Table 2) highlight the importance of performing microeconomic evaluations to cover perspectives of different decision makers when dealing with OH issues [37, 39, 42]. *Hutton* suggests that one potential way to do this is to perform both these analyses simultaneously, to provide each decision maker with the outcome estimates they need in their decision making (e.g. cost-utility outcome measures for the Health Minister and cost-benefit outcome measures for the Agricultural or Environmental Ministers) [39].

Out of the 73 studies that underwent data extraction, 52 (72%) and 45 (62%) had main outcomes of non-monetary and monetary valuations respectively. Twenty-four of these included both, many of which were climate change/emissions related interventions (15/24) that included an emissions (environmental) impact and a subsequent cost/Gross Domestic Product Impact. A literature review of outcomes in OH evaluations found that around a third of included studies (31/95) had both monetary and non-monetary outcomes [43], with the majority including some form of monetary in the form of cost-benefit and cost-effectiveness analyses [43].

In the environmental literature, monetary values of health outcomes were termed as the “value for statistical life” or “value of per loss of life years” (such as 40,000 EUR per loss of life year [44]). It should be made clear when monetizing health outcomes, why such values are chosen and theoretically what they represent. Using willingness-to-pay measures that are based on Gross Domestic Product may weight life in favour of higher-income countries [39]. Willingness-to-pay approaches that are based on primary collected data (either through revealed or stated preference studies) within the setting of interest are recommended when applying monetary value to health outcomes (if feasible) [45].

Twelve studies used statistical evaluation techniques; including regression modelling techniques [46, 47], trend analysis [48, 49] and use of basic descriptive statistics [50, 51]. Robust epidemiological and outcome data are needed for statistical evaluations to accurately understand what is currently happening and to accurately predict what might happen in the future [24]. Such data can feed into mathematical model-based evaluations [24], such as that used for the economic evaluation of climate protection measures in Germany [52]. This study performed econometric evaluation of data to estimate mathematical model parameter values and then fed these into structural equations for forecasted outputs. This approach is a useful way to ensure robust parameterisation but requires accessible, relevant data.

The aforementioned study is also an example of climate change evaluation based on previous models, extending and adapting previous economic evaluations [52]. For example, the ‘GDynE’, which was previously adapted from ‘GDyn’, was linked with the ‘GTAP-Power’ database to create a computable general equilibrium model and a cost-benefit analysis [53]. Similarly, an example related to infectious disease dynamics is the application of the NAADSM (North American Animal Disease Spread Model) to Influenza emergence in Canada [54]. However, it is also mentioned that code sharing and model reuse can lead to inexperienced people using these methods in inappropriate ways, producing flawed results [42]. It is therefore key to ensure code sharing is accompanied with clear documentation on its uses, and collaboration across model creators and users occurs.

The majority of studies found (54/73) used mathematical simulations techniques, which included not only cost-benefit and cost-utility analyses (10/73), but also basic calculations, computable general equilibrium and systems dynamics models. One cost-utility study of rabies vaccination separated out potential outcome measures useful to different decision-makers; presenting estimates of monetary expenditure, DALYs averted, animal welfare and dog acceptance (the latter two being “qualitative scores”) and estimated uncertainty through probabilistic sensitivity analysis [55]. In contrast, few of the found computable general equilibrium analyses and systems dynamics models explicitly quantified uncertainty, instead running just a small number of simulations via scenario analyses [56, 57].

An online resource aimed at providing guidance for conducting economic evaluations related to environmental adaptation policies – the ECONADAPT toolbox– highlights the importance of robustness and uncertainty when evaluating climate change policy [64]. It advocates for ‘minimising regret rather than maximising utility’ [64]. Listed methods that explicitly incorporate uncertainty include robust decision making, iterative risk management, portfolio analysis and real options analysis [64]. Only the latter was found in an applied case within our review, whereby a real options analysis was used to evaluate pandemic adaptation policy [65]. Real options analysis has a similar foundation to cost-benefit/cost-effectiveness analysis, as it uses decision-tree structures. However, the nodes in this instance represent risk events [64].The emerging infectious diseases pandemic example, compares a business-as-usual scenario, a policy implementation scenario and a “continuation value” scenario (business-as-usual plus value of waiting) for different levels of damages [65]. This essentially allows for the modelling of when best to invest. However, this method is potentially input and resource heavy, especially when applied to the dynamics of a OH issue over time [64, 65].

Multi-criteria decision analysis is also presented within ECONADAPT, allowing for flexibility in decision criteria and the ability to include uncertainty as a decision criteria in itself [64]. Of the 14 studies found that used index creation as a form of evaluating OH interventions, six studies utilised this approach (the other eight index-based studies focused on ‘One Health-ness’ [13]). One example is the application of multi-criteria decision analysis to Lyme disease management strategies [66]. In this example, multiple interventions were ranked based on public health, animal and environmental, social impact, economic, strategic, operational and surveillance criteria [66]. However, in order to rank policies based on criteria such as “reduction on incidence of human cases” and “impact on cost to public sector” [66], the outputs of epidemiological and health economic models are needed. Such methods can also incorporate other important outcome measures such as equity [40].

The multi-criteria decision analysis process requires substantial stakeholder participation. Previous reviews highlight the need to involve stakeholders (from every level) throughout the evaluation processes, across many different methodological approaches [15, 24, 41, 42]. One review highlighted the importance of thinking of end user and involving stakeholders in the model development process, however it found that only 23% of the 184 studies reported a consultation of stakeholders [41].

Most studies found had a time-horizon less than 50 years (15/73 studies used less than or equal to 1 year, 35/73 between 10 and 50 years). Those that had longer time horizons (14/73 studies had 50+ years^1^) tended to be those which had a macroeconomic approach and used computable general equilibrium, cost-benefit or multi-criteria decision approaches. The majority of papers that had a time horizon of less than ten years were using statistical evaluations of data or collecting information to create indices. Such studies are generally limited by the time period of data collection. However, further use of time-series data across a longer time horizon would be useful in understanding and forecasting the longer-term burden associated with the longer-term impact of interventions targeting such issues [67].

A discount rate is a factor by which you multiply outcomes to value future benefits/costs in today’s ‘values, accounting for factors such as time preference [68]. Debate is ongoing about the use of discount rates where many effects are seen far into the future (for example in the case of vaccination [68]). Reviewed studies used discount rates ranging from 1% to 9 % [69, 70], with some utilising a social discount rate vary over time [25, 62] and some testing multiple discount rates through scenario analyses [70–72]. National and international reports on discounting practices for the intended outcomes of interest should be sought, with sensitivity analyses being used to assess the effect of discount rate assumptions [73].

### (ii) Evaluating Antibiotic-resistance Interventions

Previous guidance has focused on recommending the first steps of policy implementation evaluation [42], as discussed in the above sections. Applying these to the hypothetical case study resulted in the following:

1. Research aim: To estimate the cross-sectoral impact of reduced antibiotic usage in agriculture and human healthcare. This could be further defined by specific antibiotic susceptibility profiles, bacterial species, infection types, human population, animal population, environmental scope and geographical scope. Other clarifications are needed such as whether multi-resistant infections or co-infections are allowed to occur [23].
2. System: A previously published generic systems map of the ABR problem has been adapted to highlight the economic agents described in Table 2 to give Figure 1 [35].

**Figure 1.**
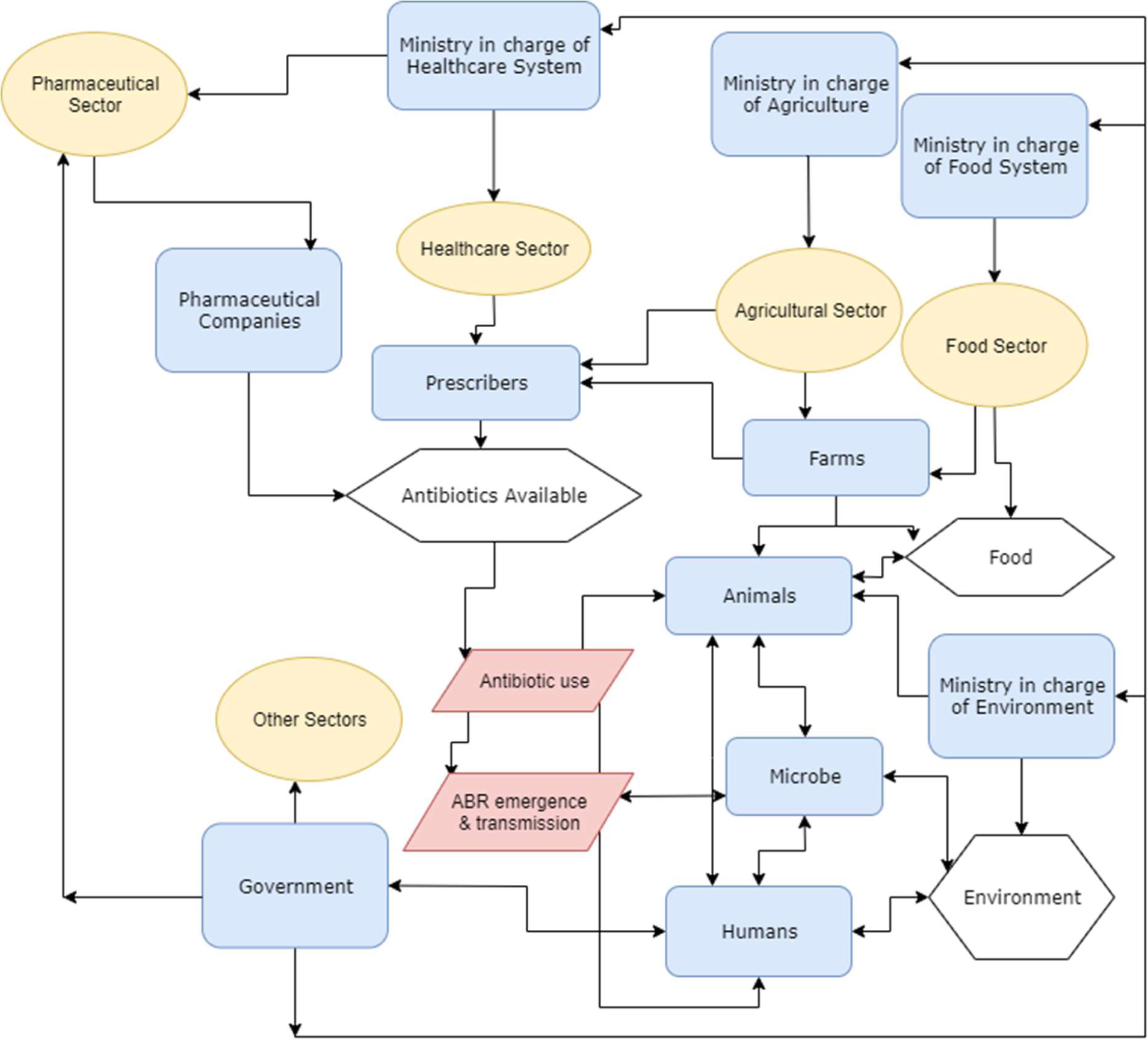
The System under Evaluation for Cross-Sectoral Antibiotic Stewardship Interventions. This system is adapted from Ruegg et al [35]. Ovals represent sectors, boxes represent agents, hexagons represent resources and parallelograms represent actions related to antibiotic stewardship. Connecting lines represent potential relationships. ‘Ministry’ may be multiple ministries in reality (for example, food system may include commerce and additional governmental offices). ABR: antibiotic resistance.
3. Defining evaluation outcome estimates that feed into decision makers’ objective functions: Figure 1 shows that there are many agents within the defined system, whilst Table 2 highlights what the objective function of such agents could be, combining these gives the potential evaluation outcome measures to be estimated (linked to specific decision makers) depicted in Figure 2.

**Figure 2.**
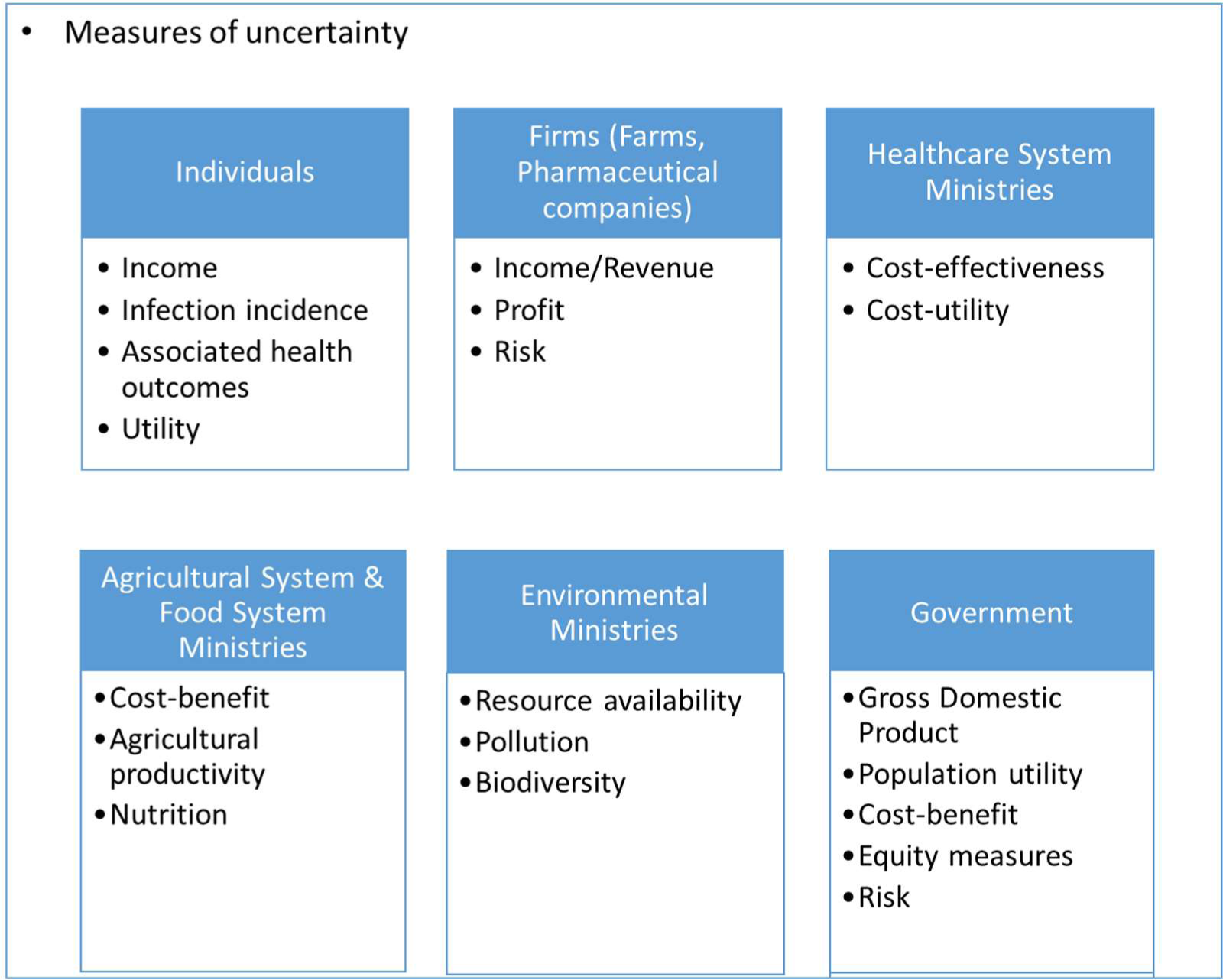
Potential Impact Evaluation Outcome Measures within the Cast Study. Bullet points represent potential evaluation measures, other box text presents related economic agents.

Though animals and bacteria are actors that can adapt to specific environments (e.g. change eating habits or gene mutation rates based on environmental status), discussions of animal behaviour and bacterial cognisance are not directly relevant to this case and are beyond the scope of this paper.

Pharmaceutical company profit calculations are subject to business sensitive information, however discussions of antibiotic research and development policy and the pharmaceutical industry can be found elsewhere [74, 75].

Prescribers may not have an objective function aligned to the individual consumer (patient), however, for the first iteration of the multi-level conceptual model, it is assumed these are aligned with the Ministry/ies in charge of healthcare, and investigated through policy implementation scenarios. ‘Prescriber’ does not necessarily equate to the traditional pharmacist or veterinarian role, but can be any intermediary within the system through which antibiotics could be purchased, such as through drug shops in low- and middle-income countries. As Figure 2 is a generic system, the flows and magnitude between outlined the different decision makers and resources would differ according to setting.

#### Methodological perspective

One potential way to produce most of the aforementioned outcome estimates is to link multiple compartmental modelling techniques. The outputs of these can then be weighted to give a composite ranking of policy options using multi-criteria decision analysis methods (using stakeholders) [66]. The suggested compartments have been outlined in Figure 3. The case study highlights disease in human and animal (livestock) compartments as this relates to the chosen case study, but the same process could be applied to include companion animals, aquaculture, and others.

**Figure 3.**
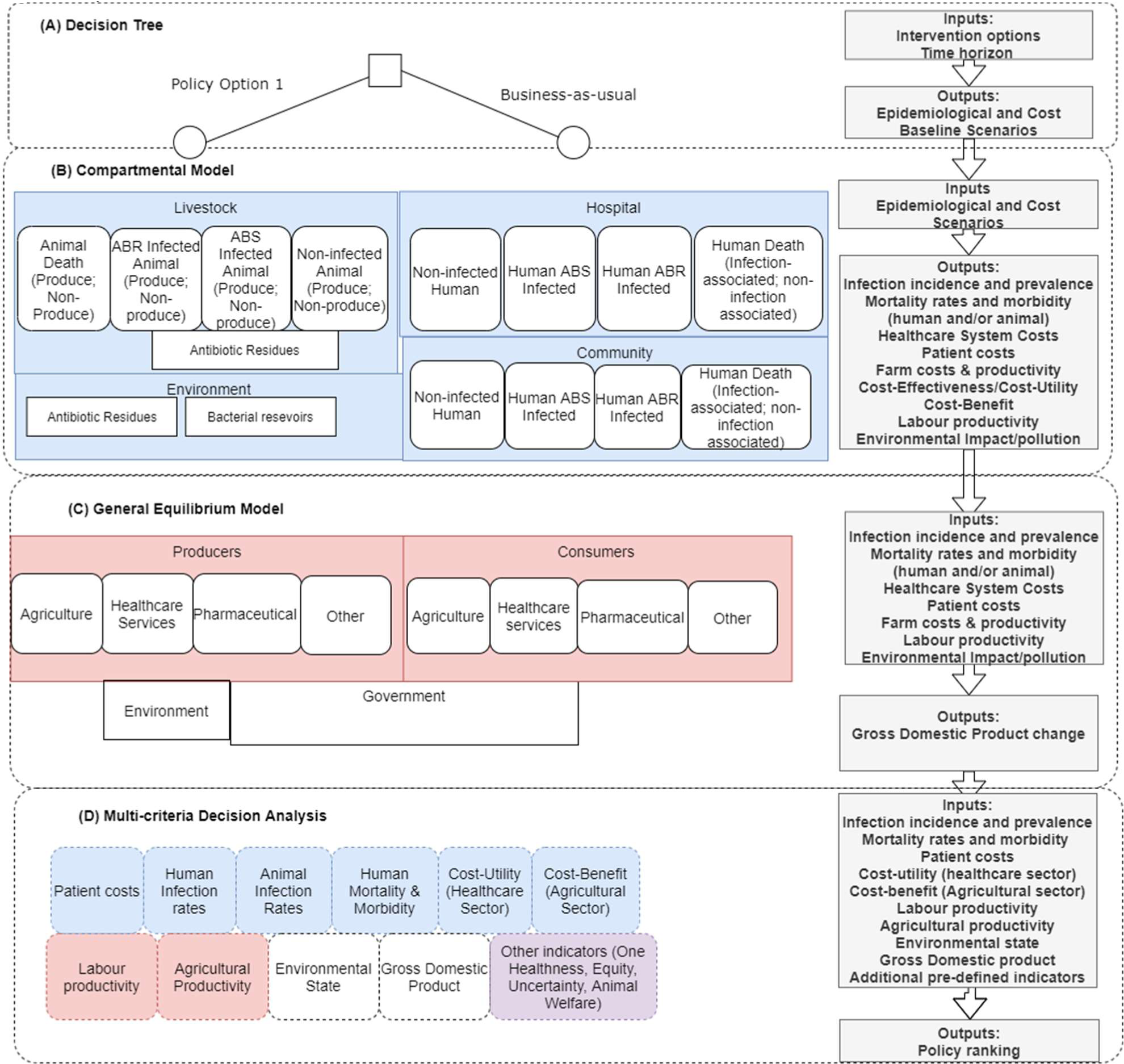
A Conceptual Multi-level Model for Evaluating Cross-Sectoral Antibiotic Stewardship Interventions. White boxes represent health states or sector states. Segments (A) to (D) represent the model method. Shaded boxes represent settings in (A) – (C) and respective model results in (D). Transitions can occur between white boxes within each segment (including across setting), such as from animal ABS carrier to Susceptible Human Carrier within (A), but these lines have not been added for visual simplicity. Boxes ‘antibiotic residues’ and ‘bacterial reservoirs’ are food and environmental states respectively. Abbreviations: ABR – antibiotic resistance, ABS – antibiotic susceptible,

From drawing out an initial conceptual evaluation in Figure 3, we can see which parameters are needed for decision tree (A) and compartmental model (B). Statistical data analyses could be used to understand factors such as: trends in resistance levels, demography of the populations of interest, specific factor productivity levels and outcomes associated with antibiotic susceptible and resistant infections (across humans and animals). The selection of models depicted in Figure 3 (B) and (C) allows for the outputs of one model to become the inputs of the next. The compartmental model structure in Figure 3 (B) also allows for the attachment of monetary benefits to the human healthcare system (such as applying a monetary value per life years lost), allowing for an aggregation of monetary costs and benefits across human healthcare and livestock systems, giving a cross-sectoral cost-benefit estimate. Humans or animals may transition across any of the health states within each segment (Figure 3 (B)). Whilst for the general equilibrium model, the compartments are representative agent states for which activity (transitions of economic inputs and outputs) occur by utilising functional definitions of agent behaviour.

Given that the implementation of ABR-related policy is over multiple years and effects multiple cohorts, the evaluation model would be multi-cohort (i.e. not just follow one “average” cohort of humans and animals over time). This could extended using microsimulation methods (i.e. tracking individuals through the compartments) [23]. The models can also attempt track distribution of outcomes across appropriate population groups (e.g. through sub-group analyses). Finally, each sub-model should calculate uncertainty explicitly, and also describe which type of uncertainty is being calculated (for example parameter uncertainty versus structural uncertainty [76]). This allows for equity and uncertainty measures to be available for the multi-criteria decision analysis stage of evaluation. Across the model a longer time-horizon (e.g. of 100 years) is recommended to account for individual lifetime horizons, however outcomes can be estimated at lesser time points if that’s what is needed according to the decision maker’s objective functions. Appropriate discount rates should be used accordingly [77], with nationally-preferred rates applied if available.

To build the whole multi-level model (conceptualised in Figure 3) through an individual project is likely unfeasible, however, the proposed structure allows for a compartmentalised build up and integration of knowledge accumulation and parameterisation overtime. Utilising the model-sharing and –adaptation approach seen in the climate change literature, this could be feasible to complete in the medium term. Once built, the model could also be expanded to allow for more complex structures and feedback mechanisms to be integrated, such as those in system dynamics models. A code-sharing website has been set up to help facilitate this work: https://zenodo.org/communities/amr-evaluation/.

## Discussion

This paper provides the first conceptual impact evaluation model of ABR-related interventions across the OH system, outlining the different stakeholders’ perspectives that should be considered when conducting such evaluations. This is done through a narrative review of the cross-sectoral intervention evaluation literature. It highlights the approach and modelling methods that could be used to carry out such an evaluation; providing a resource for future ABR-economic evaluations.

We propose ABR-related intervention evaluations take individual, microeconomic (farm and sectoral) and macroeconomic (government) evaluation perspectives across, with uncertainty estimates included for each perspective-related outcome measure. However, given the results of the literature review, it is recommended that stakeholders be involved at the onset of the development of the evaluation model. This may be informally, or researchers could engage in more formal processes of participatory modelling [78]. Stakeholder engagement is needed to define what social justice grounding will be used and to establish objective functions, to state which policy options are feasible in practice, quantify stakeholder willingness-to-pay for outcomes (e.g. health gains) and weighting of importance for intended estimated outcomes (through processes such as multi-criteria decision analysis). After this, compartmental models can be built, linked and shared following the pathway of animals and humans from colonization from bacteria, to being factors within the economy at large. Outputs of the multi-level model and qualitative work can then feed into a multi-criteria decision analysis which allows for the ranking of proposed policies taking into multi-attribute utility functions. If the proposed evaluation outcome measures and model methods are not found to be preferred after initial stakeholder involvement, the literature review’s narrative results within this study can still act as a resource for alternative methods used in other cross-sectoral intervention impact evaluations.

This study is not without limitations. The literature review was done in a structured and not systematic way, which allowed for targeted searches relating to relevant areas (such as climate change literature [17]) for a narrative discussion of methods, but subsequently meant that the relative OH perspective proportions (such as 84% including an environmental impact) cannot be taken as robustly representative of the total research space. Additionally, a generic conceptual evaluation model was applied based on literature and the authors’ judgement. A more formal process of stakeholder involvement is recommended for future direct applications of the conceptual model. Finally, the conceptual model attempts to simplify the modelling procedures, however, a lot of data, time and monetary resources are needed to apply the proposed model in practice.

In conclusion, compartmental modelling (utilising outputs from statistical analyses of data and inputs from stakeholders) teamed with multi-criteria decision analysis can allow for ABR-related interventions to be quantitatively evaluated in a way that maximises evaluation use for decision-makers across the OH system. A more open, collaborative and strategic modelling research agenda is called for, to avoid duplication and allow for more complex, complete models to be built through the linkage of sub-model information in the future.

## Data Availability

The data used in this study are in the form of literature data extraction. The data extraction table is available in the supplementary material of this paper.

## Abbreviations

ABR: Antibiotic Resistance
NEOH: Network for Evaluation of One Health
OH: One Health

## Acknowledgements

The authors would like thank Dr. Laura Cornelsen for her input in conveying the messages of this research. The authors would also like to thank Prof Jonathan Rushton for his help in understanding field of antibiotic resistance and One Health.

## Funding

This research was funded by, and is a contribution to, the CGIAR Research Program on Agriculture for Nutrition and Health (A4NH). We thank all donors and organizations which globally support its work through their contributions to the CGIAR system.

The opinions expressed here belong to the authors, and do not necessarily reflect those of A4NH or CGIAR.

## Conflicts of Interest

The authors declare they have no conflicts of interest.

## Footnotes

1 The remainder of the 73 did not state a time horizon or it was not applicable.

## Notes

### Competing Interest Statement

The authors have declared no competing interest.

